# Genomic wastewater surveillance of human and animal influenza A viruses in California during the 2024-2025 flu season

**DOI:** 10.64898/2026.06.10.26355323

**Authors:** Audrey Liwen Wang, Alexandra Lamtyugina, Minxi Jiang, Alexander T. Yu, Chunye Lu, Debra Wadford, Elisabeth Burnor, Lenore Pipes, Rose Kantor, Kara L. Nelson

**Affiliations:** Department of Civil and Environmental Engineering, University of California, Berkeley, CA, USA; Pacific Biosciences Research Center, University of Hawai’i at Mānoa, Honolulu, HI, USA; California Department of Public Health, Center for Infectious Diseases, Richmond, CA, USA; California Department of Public Health, Center for Laboratory Sciences, Richmond, CA, USA; Physical and Life Sciences Directorate, Lawrence Livermore National Laboratory, Livermore, CA, USA

## Abstract

**Background:** Wastewater genomic surveillance provides an opportunity to detect human and animal influenza A virus (IAV). We aimed to implement an IAV genomic surveillance framework agnostic to subtype, which enables recovery of IAV from multiple hosts and estimation of proportions across subtypes.

**Methods:** We conducted IAV genomic surveillance in wastewater during the 2024-2025 flu season at multiple sites in California and compared these data with available human clinical IAV sequences and test positivity. We applied a custom whole-genome, multi-host IAV probe enrichment panel and adapted our custom expectation-maximization (EM) algorithm to deconvolute IAV mixtures in wastewater and infer subtype relative abundances. Absolute IAV concentrations were quantified using RT-PCR-based assays. H5N1 wastewater and clinical sequences were further characterized by constructing a whole-genome maximum-likelihood phylogenetic tree. Finally, we performed variant analysis to examine amino acid substitutions detected in wastewater.

**Findings:** Our IAV probe enrichment method and EM algorithm successfully enriched all eight segments of three circulating IAV subtypes and accurately estimated subclade relative abundances for mixed IAV samples. Seasonal human H1N1pdm09 and H3N2 were detected throughout the study period from both wastewater and clinical sequencing data, with H1N1 subclades 6B.1A.5a.2a.1 and 6B.1A.5a.2a co-circulating, and H3N2 dominated by subclade 3C.2a1b.2a.2a.3a.1. Wastewater surveillance consistently detected H5N1 clade 2.3.4.4b across three monitored wastewater sites, while clinical H5N1 detections, from anywhere in CA, were sporadic and rare. Whole-genome phylogenetic analysis revealed that wastewater H5N1 sequences clustered with reference sequences associated with dairy cow and avian infections, while all human clinical H5N1 sequences clustered exclusively with reference sequences associated with dairy cow infections. Amino acid substitutions were identified across viral segments, and no mutations associated with mammalian adaptation were observed from wastewater samples.

**Interpretation:** When IAV concentrations were dominated by seasonal human subtypes rather than H5N1, subtype patterns aligned between wastewater and clinical data. While sequencing IAV in wastewater was unable to distinguish if H5N1 detections were due to human or animal infections, it was able to provide clade-level information about H5N1 found in wastewater that could be useful in the future. Wastewater genomic surveillance can complement clinical surveillance, increasing ability to detect all circulating IAV subtypes and enhancing public health preparedness from a One Health perspective.

**Funding:** UCOP Lab Fees CRT Award (L22CR4507) and NIH R00 Award (4R00GM144747-03)

**Research in context:** *Evidence before this study:* Sequencing IAV in wastewater has been shown to detect circulating subtypes from human and animal hosts, with potential to improve public health response, enhance surveillance for novel pandemic threats, and inform vaccine design. However, methodological improvements are needed for this potential to be fully realized. Targeted enrichment methods designed for whole-genome sequencing from wastewater can substantially improve genome coverage and sensitivity for low-abundance IAV. Approaches have included tiled amplicon, universal amplicon, and probe-capture enrichment. Tiled-amplicon methods provide high sensitivity and specificity but are less tolerant to sequence mismatches, and typically restrict primer design to a limited set of segments and a narrow range of subtypes. The universal amplicon approach was designed for clinical sequencing, where intact genomes from single isolates are available, and previous studies showed low recovery when applied to wastewater samples. In contrast, probe-capture enrichment tolerates mismatches and is therefore more resilient to RNA degradation and better suited to capture novel or divergent variants. However, current commercial probe-capture panels are pan-viral, which reduces sensitivity for IAV due to the high number of reads corresponding to other higher-abundance targets. The segmented nature of the IAV genome introduces additional challenges for downstream bioinformatic analysis. Existing deconvolution tools for wastewater sequencing have been developed primarily for SARS-CoV-2 and are not readily applicable to IAV. When applied to IAV, existing tools often demix only one subtype at a time, inferring clade-level relative abundance for that specific subtype. This limitation arises because different IAV subtypes require distinct reference genomes, unlike SARS-CoV-2, where a single reference can support alignment of multiple variants. Given that wastewater samples often contain multiple IAV subtypes simultaneously, it would be more useful to deconvolute clade-level relative abundances for all subtypes in parallel, underscoring the need for a more robust method tailored to the genomic complexity of IAV in wastewater.

*Added value of this study:* To our knowledge, this is the first study to apply an IAV-specific probe-capture enrichment panel to monitor IAV in wastewater, targeting all eight segments of the genome and subtypes across human, avian, dairy cattle, and other mammalian hosts. Unlike studies that prioritize either high coverage of very few subtypes or broad detection with limited genomic resolution, our study achieves a balance between capturing multiple IAV clades and maintaining reasonable coverage across segments. Additionally, we adapted a probabilistic expectation-maximization (EM) model to infer clade-level relative abundances from wastewater sequencing data. Our method includes imputation and retains fine-scale genomic differences, reducing bias when genome coverage is incomplete, which is a common scenario for low-abundance IAV in wastewater. While several bioinformatic tools have been developed to deconvolute SARS-CoV-2 mixtures in wastewater, comparable approaches have not been tailored for IAV, whose segmented genome and extensive subtype diversity pose unique challenges. Our framework fills this gap by enabling robust subclade-resolution surveillance across multiple subtypes and hosts using sequence data from wastewater samples.

*Implications of all the available evidence:* Our findings demonstrate a framework for wastewater genomic surveillance for tracking human IAV while also detecting animal-associated IAV. From a public health perspective, multi-host wastewater genomic surveillance offers two key benefits. First, it strengthens existing human IAV surveillance, because sequencing of clinical flu samples is limited and may not be representative of circulating subtypes, especially across regions with differing resources. Wastewater surveillance captures a larger and less biased sample population and has the potential to detect and track new subtypes more efficiently than clinical surveillance. Second, the ability to detect animal-associated IAV in wastewater supports pandemic preparedness by enabling monitoring for IAV of zoonotic potential and of genetic markers indicative of potentially increased human adaptation. This is especially beneficial where active surveillance of animal hosts is limited.

## 1. Introduction

Influenza A viruses (IAV) cause seasonal flu, primarily driven by H1N1pdm09 and H3N2, and pandemics that often arise from zoonotic spillover.^1^ In March 2024, the first avian H5N1 clade 2.3.4.4b was reported in U.S. dairy cattle, followed by confirmed human cases without sustained human-to-human transmission. In the second half of 2024, California reported large number of H5N1 infections in dairy cattle herds, and has accounted for more than half of the human H5N1 cases in the U.S. to date.^2^

Wastewater-based surveillance has emerged as a promising One Health tool for detecting both human and animal-associated pathogens.^3^ CDC’s National Wastewater Surveillance System monitors influenza concentrations in wastewater,^4^ complementing existing clinical surveillance systems^5,6^ that are often constrained by resources, access, and time. However, current IAV wastewater surveillance primarily relies on RT-PCR-based approaches targeting the Matrix (M) or Hemagglutinin (HA) genes, which quantify overall IAV abundance but provide limited subtype or clade resolution.^7,8^ Sequencing-based wastewater monitoring could enable detection of emerging variants or novel strains, as well as provide finer clade and even subclade classification.^9-11^

However, sequencing IAV genomes from wastewater remains challenging because IAV is present at low concentrations relative to the total microbial community. As a result, effective amplification or enrichment steps are often required prior to sequencing. Previous studies have applied methods such as tiled amplicon,^9,12-14^ universal amplicon,^11,12^ or probe-capture enrichment,^12,15-17^ to increase IAV detection sensitivity and genome recovery. Nevertheless, most tiled amplicon studies have focused more on the surface glycoprotein genes, HA and neuraminidase (NA), of human seasonal IAV like H1N1pdm09 and H3N2,^9,14^ which could overlook animal-associated IAV strains as well as information about internal segments like PA, PB1, and PB2 where mammalian adaptation and host-determination variants are often located. Low IAV recovery was observed using the universal amplicon method, due to its reliance on intact genome segments.^11,12^ Meanwhile, probe capture panels^15,16^ often target multiple viruses simultaneously, which reduces sequencing depth specifically for IAV.^18^

Additionally, unlike clinical samples, a single wastewater sample may contain multiple subtypes and strains of IAV. Bioinformatic analysis requires deconvolution to estimate strain-level abundance, but most deconvolution tools were developed for SARS-CoV-2.^19-21^ While SARS-CoV-2 has a single-genome structure, IAV possesses a segmented genome. Consequently, when these tools are applied to IAV, deconvolution is often restricted to analyzing one segment and one subtype at a time. For example, Freyja demixes H1N1 to 5a.2a.1, 6B.1A, 6B.2, etc. based on the HA segment only.^19^ Another IAV surveillance study applied LolliPop^20^ for demixing, which produced subclade abundance for H1N1 but not for H3N2.^9^ To our knowledge, no current computational tools can simultaneously deconvolute multiple IAV subtypes to a subclade level for a single sample.

To address these challenges, we implemented a custom IAV-specific whole-genome probe capture enrichment panel targeting 11 IAV subtypes across multiple hosts, enabling One Health genomic wastewater surveillance. In addition, we compared two methods, EsViritu^16^ and a custom expectation-maximization (EM) based deconvolution tool, to estimate IAV clade/subclade relative abundances from the wastewater sequencing data. EsViritu was selected because it allows multiple reference sequences as input, enabling clade-level deconvolution of whole-genome, segmented viruses such as IAV in wastewater samples. Sequencing and ddPCR data generated from wastewater samples were also compared with clinical sequencing data and test positivity. H5N1 wastewater and clinical sequences were further characterized by constructing a whole-genome maximum-likelihood phylogenetic tree. Finally, we performed variant analysis to examine amino acid substitutions detected in wastewater.

## 2. Methods

### 2.1 Wastewater sampling and nucleic acid extraction

Samples were collected weekly from three wastewater treatment plants (WWTPs) in California between November 2024 and February 2025. Samples were 24-hour composites from three sites: East Bakersfield Treatment Plant (BKERSFLD), Central Contra Costa Sanitary District (CCCSD), and Los Angeles County Sanitation Districts (LACSD). These sites represent varying population sizes and operating conditions (**Table S1**). All wastewater samples were collected in 50-mL falcon tubes in replicate, shipped on ice to the Drinking Water and Radiation Laboratory (DWRL) at the California Department of Public Health (CDPH), Richmond, and stored at 4 °C for up to five days prior to wastewater extraction.

Up to two 50 mL aliquots of wastewater from each WWTP were picked up from CDPH weekly, transported on ice to the University of California, Berkeley (UCB), and extracted on the same day. The UCB-extracted samples were used for IAV probe-capture enrichment sequencing experiments in this study and compared with wastewater and clinical results from CDPH’s routine monitoring. For extraction at UCB, 40 mL aliquots were extracted using the Promega Wizard Enviro TNA Kit (“Promega”) according to the manufacturer’s instructions with detailed procedures described previously.^22^ This kit was selected as the extraction method due to its high IAV recovery efficiency as measured by dPCR^22^ and its second-highest sequencing sensitivity in our previous benchmarking studies.^15^ Nucleic acid extracts were aliquoted and stored at -80 °C for up to one month before dPCR quantification and up to three months before library preparation and sequencing. All extracted nucleic acid samples underwent one freeze-thaw cycle. For CDPH’s routine wastewater monitoring, the KingFisher automated magnetic-bead protocol was used following their protocol (**Supplementary Methods B**) adapted from Karthikeyan et al. 2021^23^ to concentrate and extract viral nucleic acids from wastewater, followed by downstream quantification using RT-ddPCR.

### 2.2 Total IAV and H5 subtype quantification via RT-dPCR and -ddPCR

IAV M-gene concentrations, representing total IAV abundance, were quantified for all UCB-extracted wastewater samples using the QIAcuity Four Platform dPCR system (Qiagen). Detailed information about UCB’s dPCR assays and the Environmental Microbiology Minimum Information Checklist^24^ are provided in **Supplementary Methods A**. For CDPH’s routine IAV monitoring, M-gene, H5, and PMMoV concentrations were quantified by ddPCR also following their protocol (**Supplementary Methods B**).^23^ To obtain the most complete data for H5 concentrations, data from multiple laboratories were used: DWRL at CDPH, as well as partner programs (including the CDC National Wastewater Surveillance System (NWSS) wastewater testing contract and WastewaterSCAN),^25^ whose results are reported to the CDC and made publicly available through the California Wastewater Surveillance Dashboard.^8^ Primer and probe sequences for all assays are listed in **Table S2**. The ddPCR PMMoV-normalized M-gene data (**Table S5**) were also plotted alongside the PMMoV-normalized H5 data (**Table S6**) for comparison (**Figure S1)**. PMMoV normalization helps to account for wastewater dilution by stormwater. To assess consistency between extraction and quantification methods, dPCR M-gene concentrations measured by UCB were compared with ddPCR reported by CDPH (**Figure S2** and **Table S5**).

### 2.3 Influenza clinical data

Two types of clinical data from CDPH and CDC were used in this study: publicly available clinical IAV sequences and test positivity rates. All clinical sequences collected during the same period as the wastewater samples were downloaded from GISAID^26^ by filtering for sequences with the “California Department of Health Services” listed as the Originating Laboratory. Each deposited sequence corresponds to one clinical specimen, and clinical sequencing followed random sampling protocols to ensure representative population coverage. Anonymized geographic information for clinical sequences, provided by CDPH at the city- or county-level, was used to assign each sequence to the nearest sewershed based on driving distance (**Table S7**). This decision was made because restricting the data to the smaller public health jurisdictions (**Figure 1**) resulted in exclusion of 60 (out of 246) clinical samples, which would limit the representativeness of circulating strains. Test positivity data were retrieved from the California Respiratory Virus Dashboard^6^ by selecting Condition as “Flu” and Region as “Bay Area”, “Los Angeles”, and “Central California”,^27^ as these regions encompass the sewersheds for CCCSD, LACSD, and BKERSFLD, respectively.

**Figure 1.**
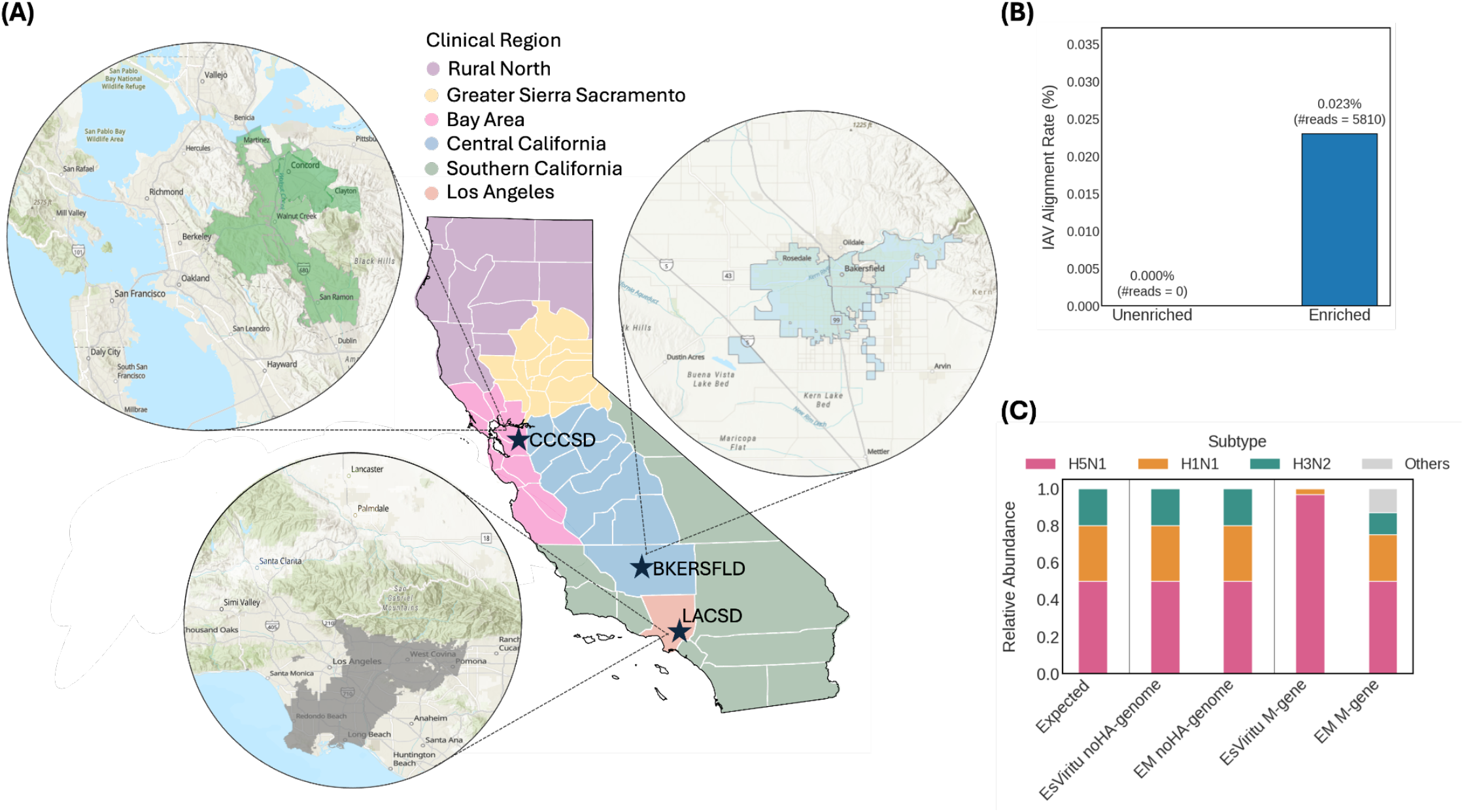
Method Overview. **(A)** Clinical influenza test positivity data represents six public health regional jurisdictions (as colored on the map): Rural North, Greater Sierra Sacramento, Bay Area, Central California, Southern California, and Los Angeles.^6^ The regional map for test positivity was retrieved from the CDPH Public Health Office^27^. Stars indicate WWTPs sampled in this study and corresponding sewershed maps are shown in circles. Wastewater data from each sewershed were compared with clinical test positivity in the corresponding region: CCCSD with the Bay Area (pink), BKERSFLD with Central California (blue), and LACSD with Los Angeles (orange). **(B)** One BKERSFLD sample was split and processed in parallel as enriched and unenriched to compare probe-capture efficiency. IAV alignment rate (%) was calculated as the number of reads aligned to the IAV reference database divided by the total number of quality-filtered reads; “#reads” denotes the count of IAV-aligned reads. **(C)** Simulated Illumina reads consisting of 50% H5N1, 30% H1N1, and 20% H3N2 (shown as “Expected”) were analyzed using two bioinformatic pipelines, EsViritu and EM, to evaluate relative abundance estimation accuracy. For each pipeline, results were compared between databases constructed from either whole-genome without HA sequences or M-gene sequences only.

### 2.4 Custom IAV probe enrichment panel and sequencing

We applied a recently designed customized IAV probe panel,^12^ which targets 11 different subtypes of IAV genomes, including H1N1, H3N1, H5N1, H1N2, H2N2, H3N2, H9N2, H7N9, H4N6, H5N6, and H5N9. Library preparation and IAV enrichment were performed following Twist Total Nucleic Acids Library Preparation EF Kit 2.0 and Twist Target Enrichment Standard Hybridization v2 with small modifications (**Supplementary Methods C**). One wastewater sample from BKERSFLD was sequenced with and without probe enrichment to assess enrichment efficiency. A synthetic DNA whole-genome IAV mixture was used as a positive control: for each virus (H5N1: EPI_ISL_19088566; H3N2: EPI_ISL_19407907; H1N1: EPI_ISL_19407925), the eight genomic segments were synthetized separately (gBlocks, IDT) and mixed at equal genome-copy concentrations to generate a synthetic whole-genome construct. The three whole-genome constructs were then combined at a 0.5:0.3:0.2 ratio at a total concentration of 10^8^ genome copies per liter (gc/L) and used as input for library preparation without any wastewater background. All libraries were stored at −20 °C for up to one month before sequencing. All samples were sequenced on one lane of NovaSeq X 10B 150PE (1.25 billion reads pairs), with an average of 30 million reads per sample.

### 2.5 Expectation-Maximization (EM) algorithm and bioinformatics analysis

All Illumina short reads from the probe-enriched wastewater samples were quality-trimmed using *fastp* (v0.24.0) to remove sequencing adapters, poly-X tails, and low-quality bases, and to discard reads shorter than 70 bp. Summary statistics for high-quality reads, including read counts, average length, and GC content, were obtained using SeqKit (v2.4.0). Quality-filtered reads were analyzed using both the EsViritu pipeline^17^ with a custom IAV genome database (see below) and a custom EM pipeline adapted from our previous SARS-CoV-2 method^28^ for estimating read counts and relative abundance of IAV clades. The EM algorithm was modified to accommodate the segmented influenza A virus genome and diverse subtypes presented in a sample. The modified implementation is available at: https://github.com/alamda/mismatch-matrix-merge, and the detailed description of the EM algorithm is provided in **Supplementary Methods D**. The positive control was evaluated by mapping reads to the corresponding spike-in reference genomes using Bowtie2 (v2.5.4), and normalized read abundances were calculated as reads per kilobase per million quality-filtered reads (RPKMF). Variant analysis was performed on each wastewater sample using reference sequences of A/California/07/2009 (H1N1), A/Darwin/9/2021 (H3N2), and A/dairy_cow/Texas/2024 (H5N1). Missense variant frequencies, representing amino acid-altering mutations, were calculated using a minimum read depth of 10 and an alternative allele frequency threshold of 0.2 (see **Supplementary Methods E** for details). A concatenated whole-genome H5N1 maximum-likelihood phylogenetic tree was constructed using H5N1 sequences downloaded from GISAID (November 2022-November 2025) to characterize our H5N1 sequences obtained from wastewater and clinical samples (see **Supplementary Methods F** for details).

During inspection of read counts across segments, we identified contamination from HA tiled amplicons originating from previous spike-in experiments. This contamination was confirmed by the unusually high proportion of HA reads (**Figure S4A**) and by the characteristic tiled-amplicon coverage pattern observed across the HA segment (**Figure S4B**). Attempts to decontaminate the HA reads were unsuccessful, as the signal-to-noise ratio was too low to confidently recover true probe-enriched reads. To mitigate this issue, both EsViritu and our EM deconvolution tool estimated clade relative abundances using the remaining seven segments, and a custom IAV reference database was constructed with all HA segment sequences excluded. This approach allowed us to confidently identify IAV clades present in wastewater and estimate their relative abundances. Notably, the positive-control IAV gBlocks exhibited uniform read distribution across segments (**Figure S4A**) due to their high input concentration relative to the HA tiled-amplicon contamination, whereas probe-enriched wastewater samples contained much lower IAV concentrations, making them more susceptible to contamination.

IAV reference sequences for building the custom IAV reference database were downloaded from GISAID (2021-2025, USA),^23^ filtered to retain only complete genomes. To avoid false alignments due to carryover from prior tiled-amplicon contamination, HA segment sequences were removed from the database before analysis. The highly conserved 5’ and 3’ untranslated regions (UTRs) were trimmed from all reference sequences to prevent misalignment across influenza strains and genomic segments. Identical seven-segment genomes were collapsed at 100% identity, resulting in a final database of 100,003 unique IAV isolates. This database was used for both the EsViritu and EM pipelines.

### 2.6 Data analysis

All statistical analyses were conducted using the Python package scipy.stats, with significance determined at a 95% confidence level (p < 0.05). Data analysis and visualization were performed using custom Python and R scripts. The equations for calculating IAV alignment rate (**Eq.1**), mean depth (**Eq.2**), averaged segment mean depth (**Eq.4**), coverage breadth (**Eq.5**), reads per kilobase per million quality-filtered reads (RPKMF) (**Eq.6**), and the final IAV gene-copies per liter of wastewater (**Eq.7**) are summarized in **Supplementary Methods G**.

## 3. Results

### 3.1 IAV probe capture enrichment and EM-algorithm for estimating relative abundance of clades

The probe capture enrichment efficiency was evaluated using a single wastewater sample collected on 12/19/2024 from BKERSFLD. In this sample, dPCR for the IAV M-gene detected two positive partitions out of 25,468 valid partitions, corresponding to an estimated IAV concentration of 540 gc/L-WW (**Eq.7**). Without enrichment, zero reads aligned to IAV, while enrichment yielded 5810 IAV reads out of 25.25 million reads (**Figure 1B**). Despite probe enrichment, the alignment rate remained low (0.023%), likely because of the low initial IAV concentration in the sample.

To further assess potential bias introduced by the probe panel when capturing all segments of the multiple subtypes expected to be found in wastewater, a synthetic DNA whole-genome IAV mixture was created as a positive control (see Methods). After probe-capture enrichment, the segment-level alignment rate ranged from 7.2% (NS) to 15.6% (PB2) and 99% of all reads aligned to IAV genomes (**Figure S4A**). The inferred subtype relative abundances, based on normalized reads (RPKMF, **Eq.6**) averaged across segments, were H5N1:H3N2:H1N1 = 0.583:0.25:0.167, which closely matched the expected mixture of 0.5:0.3:0.2 (**Table S10**).

Accurate estimation of viral relative abundances from sequencing data is crucial for understanding the dynamics of clades over time. We first benchmarked the relative abundances of clades estimated by two tools, EsViritu with a curated IAV database and our custom EM tool, using *in silico* simulated Illumina data (1 million reads) of IAV mixture with a proportion of H5N1:H3N2:H1N1 set at 0.5:0.3:0.2. When relative abundances were estimated using the whole-genome without HA reference database, both EsViritu and our EM tool produced accurate estimates, deviating from the expected proportions by less than 1% (**Figure 1C**). However, when estimation relied on only a single segment (Matrix gene), EsViritu exhibited a strong bias toward H5N1 reads (>0.96), whereas the EM tool more accurately reflected the expected proportions (0.499:0.251:0.119) (**Figure 1C**).

This bias was also observed when both tools were applied to the real wastewater samples. EsViritu also showed large proportions of H5N1 2.3.4.4b reads predominated across almost all samples (**Figure S5**), while the estimates generated by the EM tool ranged from 0 % to 60.6% (**Figure 2A**). Given its increased accuracy, we continued forward with the EM method.

**Figure 2.**
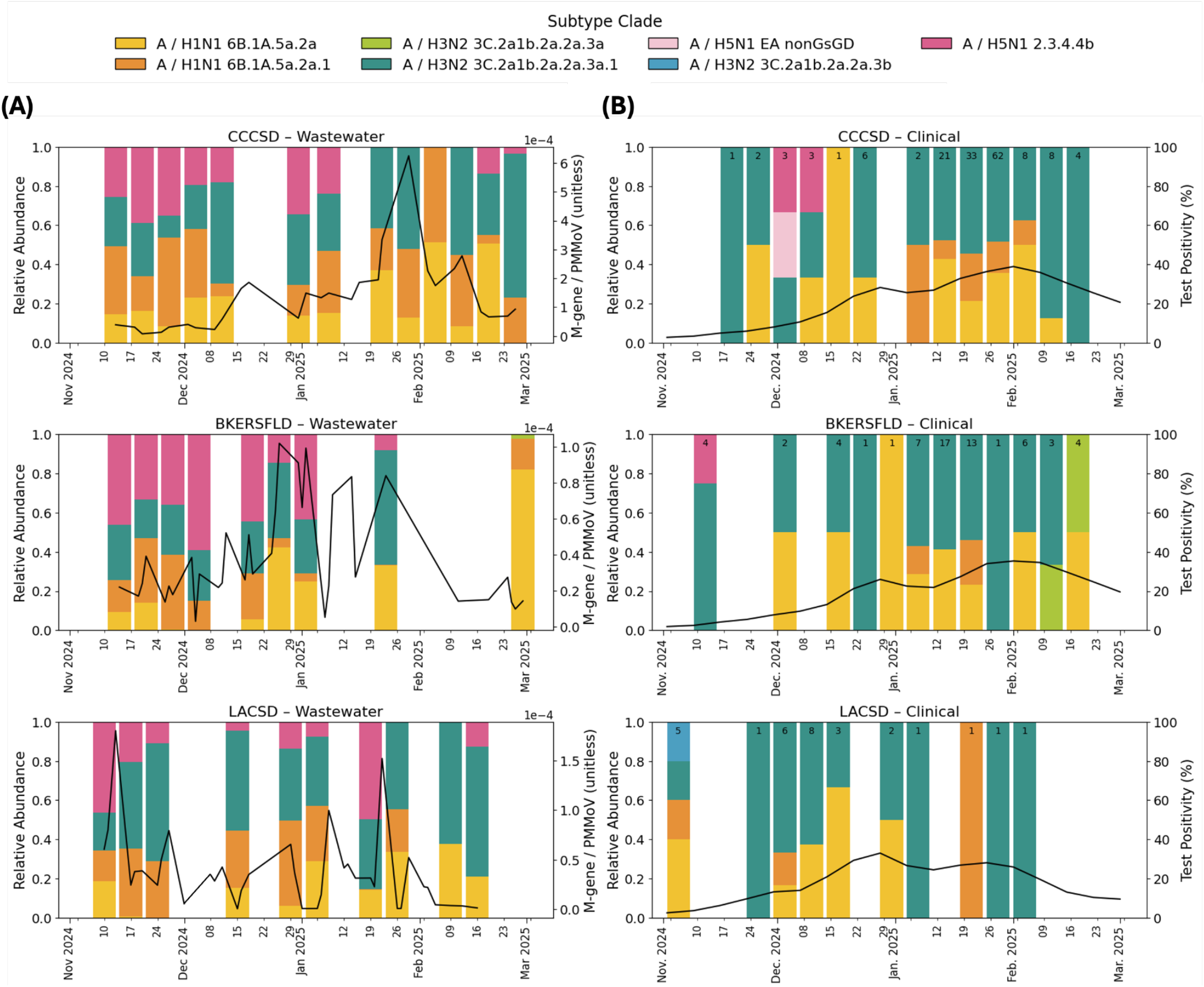
Comparison of circulating IAV subtypes (H1N1, H3N2, and H5N1) between wastewater and clinical genomic surveillance. **(A)** Relative abundance of IAV subclades across three California wastewater treatment plants from top to bottom: CCCSD (n=13), BKERSFLD (n=9), and LACSD (n=10). Each stacked bar represents a weekly sequenced wastewater sample processed by IAV probe capture enrichment panel, with colors indicating different IAV subclades (see legend). Relative abundance was estimated using the EM algorithm as described in **Supplementary Methods D**. Black line indicates ddPCR concentration of PMMoV-normalized IAV M-gene (unitless)^9^ across the sampling period. **(B)** Relative abundance of clinical sequences for the same timeframe as wastewater samples across the three WWTP catchment areas. Raw data of relative abundance are reported in **Table S8**. A total of 246 clinical sequences were included (n_CCCSD_=154; n_BKERSFLD_=63; n_LACSD_=29). Relative abundance was calculated as the proportion of clinical sequence count assigned to each subclade per week. Each stacked bar represents weekly clinical data, with the number of sequences indicated at the top of each bar. Instances of relative abundance equal to 1 occurred when only one clinical sequence was obtained from that week. Black line represents the influenza test positivity (%) in the Bay Area, Central California, and Los Angeles from top to bottom,^6^ whereas clinical sequencing samples were associated with the closest WWTP (**Table S7**).

### 3.2 IAV profiles captured by wastewater and clinical sequencing

Samples were collected weekly from three wastewater treatment plants between November 2024 and February 2025, followed by probe-capture enrichment and sequencing. From both wastewater and clinical sequencing, three IAV subtypes were detected: H1N1pdm09, H3N2, and H5N1. The seasonal subtypes associated with humans, H1N1pdm09 and H3N2, were predominant for most of the study period (**Figure 2**). For H1N1pdm09, clades 6B.1A.5a.2a.1 (orange) and 6B.1A.5a.2a (yellow) co-circulated, and their proportions varied by region and over time. H3N2 circulation was dominated by a single subclade 3C.2a1b.2a.2a.3a.1 (dark green), accounting for 99-100% of H3N2 wastewater reads and clinical sequences throughout most of the surveillance period across all sites. In the BKERSFLD site, subclade 3C.2a1b.2a.2a.3a was detected in clinical sequences during the second and third weeks of February 2025 (**Figure 2B**, light green), when wastewater samples were not collected at this site due to operational constraints. This strain was detected in BKERSFLD in wastewater in early March 2025, when sampling resumed (**Figure 2A**, light green). One clinical sequence assigned to H3N2 subclade 3C.2a1b.2a.2a.3b (blue) was detected in the LA region but was not observed in any of the wastewater samples.

H5N1 clade 2.3.4.4b was consistently detected in wastewater sequencing data across all three treatment plants (**Figure 2A**, dark pink). In contrast, H5N1 detections in clinical specimens were sporadic and rare, with only isolated instances of clade 2.3.4.4b identified and assigned by driving distance to BKERSFLD and CCCSD (**Figure 2B**, dark pink). An H5N1 Eurasian lineage (EA; light pink) was detected clinically but not in wastewater.

### 3.3 Wastewater IAV peaks dominated by H5N1 were not accompanied by corresponding clinical peaks

The overall IAV levels are represented by ddPCR PMMoV-normalized M-gene data for wastewater (**Figure 2A**, black line) and test positivity for clinical cases (**Figure 2B**, black line). Clinical test positivity (**Figure 2B**, black line) generally synchronized trends across three locations, with bimodal peaks occurring consistently in late December and late January. However, wastewater peaks (**Figure 2A**, black line) occurred at different times across locations. Most notably for example, LACSD showed an early peak in mid-November not seen at other sites. To identify the potential contributors to wastewater IAV peaks, we compared ddPCR H5 concentrations with M-gene concentrations (**Figure S1**). For LACSD, the early increase in IAV signal was largely driven by H5N1; this was true in BKERSFLD in late December as well, though to a lesser extent (**Figure S1B** and **Figure S1E)**. The H5N1-dominated peak for LACSD wastewater in mid-November did not have a corresponding peak observed in clinical data (**Figure 2**). Wastewater peaks that were not dominated by H5N1, for instance at CCCSD, showed a better temporal alignment with clinical peaks (**Figure 2**). Additionally, during the highest wastewater peak at CCCSD (mid-January to early-February, 2025), and when the sample size for clinical sequences was also large (n = 8-62), the relative abundance pattern of H1N1 clades (orange and yellow) and H3N2 (dark green) were in good agreement.

### 3.4 Phylogenetic host characterization of wastewater and clinical H5N1 sequences

To determine the most likely host origins of the H5N1 from wastewater and clinical sequences, we constructed a whole-genome maximum-likelihood phylogenetic tree using GISAID H5N1 sequences collected between November 2022 and November 2025, including sequences from four hosts: human, avian, dairy cow, and swine (**Figure 3, Supplementary Methods F**). Among available submitted reference sequences, most belonged to clade 2.3.4.4b and were isolated from avian hosts. Clades 2.3.2.1a, 2.3.2.1e, EA_nonGsGD, and Am_nonGsGD formed distinct clusters in reference sequences, and many human-associated H5N1 sequences were found within clade 2.3.2.1e, from samples collected in Cambodia/India and not associated with the 2025 dairy cow outbreak in California. All H5N1 reads recovered from California wastewater in this study were assigned to clade 2.3.4.4b. The majority were affiliated with dairy cow-associated strains (**Figure 3**, red rectangle) and were detected in 25 of 32 samples. A smaller number of reads were associated with avian strains (**Figure 3**, red rectangle), which occurred in six samples (two from LACSD and four from BKERSFLD). All human clinical H5N1 sequences (submitted by CDPH to GISAID, n=9) clustered with strains isolated from dairy cows (**Figure 3**, purple circle), and eight of them were classified as clade 2.3.4.4b, while one was classified as EA_nonGsGD.

**Figure 3.**
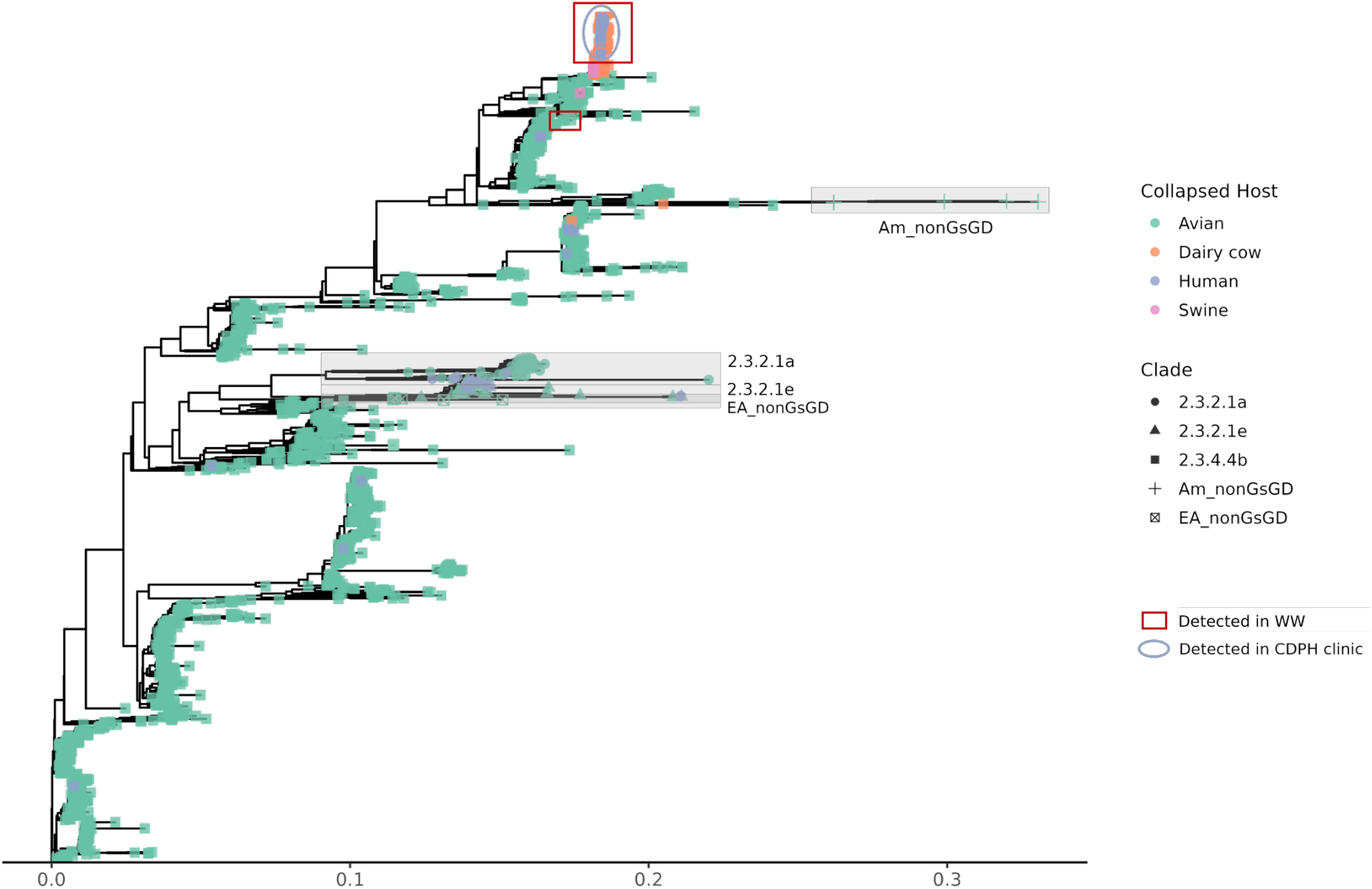
Concatenated whole-genome maximum-likelihood phylogenetic tree of H5N1. All H5N1 sequences collected between November 2022 and November 2025 (n_total_=2046: n_avian_=1868 ; n_dairy-cow_=126; n_human_=47; n_swine_=5) and their associated metadata were downloaded from GISAID. Sequences were quality-filtered and trimmed. Segments from each genome were first concatenated and aligned using MAFFT, and a maximum-likelihood tree was then constructed using RAxML. The nodes were colored by collapsed hosts and shaped by clades. The isolates that were detected in wastewater samples were highlighted with red rectangles, and those detected in our clinical samples were highlighted in purple circles.

We did not attempt to infer host abundance for the reads recovered from wastewater, as only a few nucleotide variants differentiate cattle- and avian-derived H5N1 2.3.4.4b sequences. Determining the prevalence of reads originating from either host would require coverage across these specific signature regions. Because of the similarity in 2.3.4.4b sequences across host species and the overall paucity of isolates in GISAID for which to compare, association of H5N1 in wastewater samples with available dairy cow or avian isolates does not mean that those animals were the definitive sources of H5N1 in those wastewater.

### 3.5 Coverage and variant analysis of wastewater sequences

Sequencing depth and breadth varied across IAV segments. Shorter internal segments like MP, NP, and NS exhibited more uniform read distribution across segments compared with the longer segments PB1, PB2, and PA (**Figure 4A**). For all three subtypes, the MP segment showed the highest average mean depth compared to the other segments (**Figure 4A, Table S11**, H1N1_MP_ = 15.6×, H3N2_MP_ = 17.8×, and H5N1_MP_ = 12.4×). The segments with the highest median coverage breadth for each subtype were also the shorter internal segments: MP for H1N1 (39.2%), NS for H3N2 (37%), and NP for H5N1 (28%) (**Figure S6, Table S12**).

**Figure 4.**
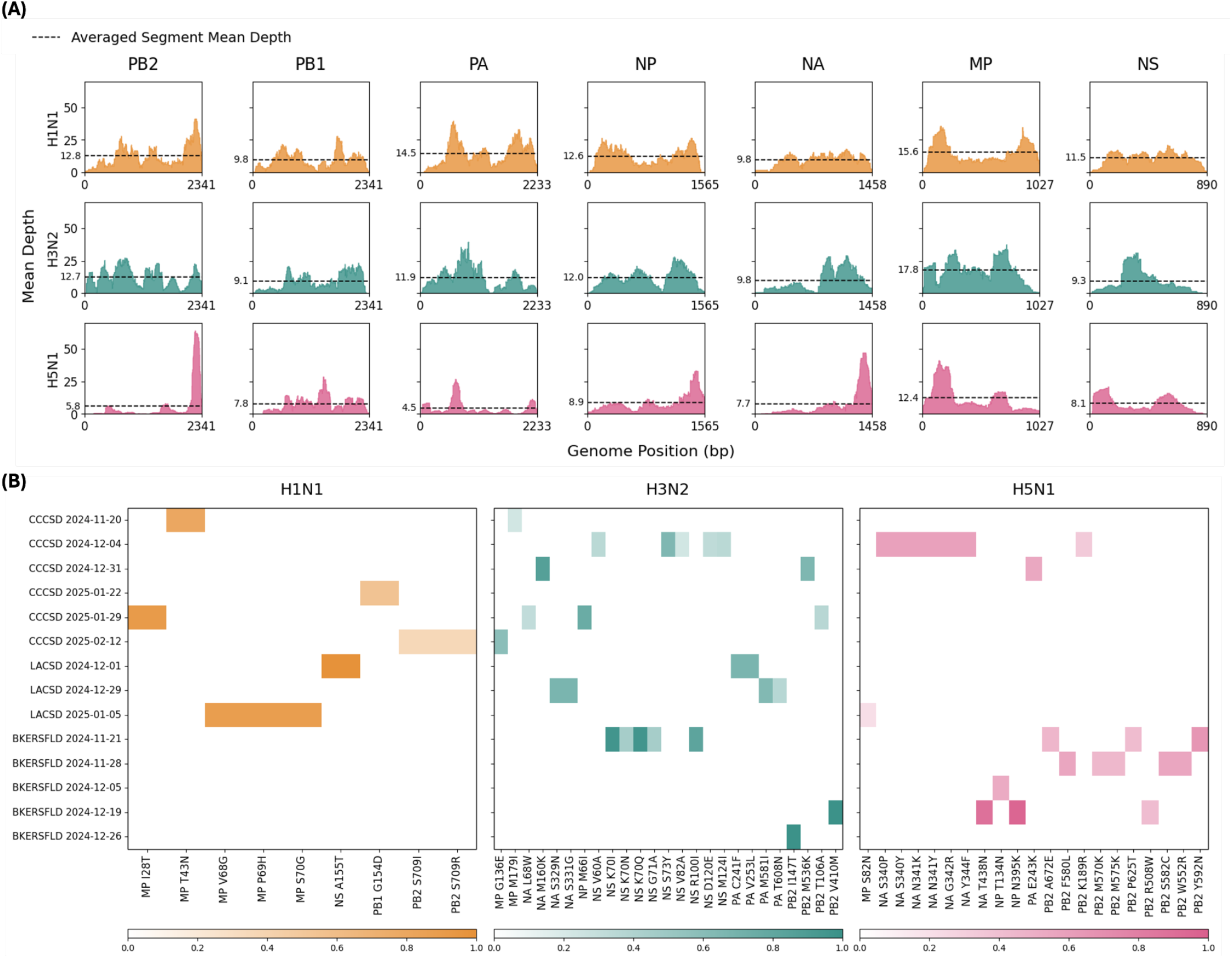
Read coverage analysis of influenza virus genome segments in wastewater surveillance across three dominant subtypes. **(A)** Histograms of mean depth (mean read count) at each genomic position (x-axis), averaged across all samples. Dashed lines indicate the average read depth across each segment, averaged across all samples. Subtypes are represented by orange (H1N1), dark green (H3N2), and dark pink (H5N1). **(B)** Heatmaps showing missense mutations (x-axis) detected in individual samples (y-axis). Color indicates the allele frequency (0-1) as a fraction of read depth at each position. Abbreviations: PB2, Polymerase Basic Protein 2; PB1, Polymerase Basic Protein 1; PA, Polymerase Acidic Protein; NP, Nucleoprotein; NA, Neuraminidase; MP, Matrix Protein; NS, Nonstructural Protein.

All probe-enriched samples contained reads aligned to IAV, ranging from 94 reads (LACSD 2/2/2025) to 11,812 reads (BKERSFLD 12/12/2024) (**Table S13**). Across all samples, the BKERSFLD sample collected on 12/12/2024, which had the ddPCR concentration of 4.13×10^4^ gc/L-WW IAV M-gene, exhibited the highest segment mean depth (161.45×), corresponding to the H5N1 MP segment. This was also the only sample with 100% MP segment coverage breadth observed across all three subtypes (**Table S11-S12, Figure S7**).

Variant analysis was performed on each wastewater sample against a single reference sequence for each subtype (see Methods) and missense variant frequencies, representing amino acid-altering mutations, were calculated (**Figure 4B**). For H1N1, amino acid substitutions (AAS) were detected only in the internal segments (MP, NS, PB1, and PB2) and were not observed in BKERSFLD. H3N2 showed AAS substitutions across multiple segments, with the NS segment accumulating more substitutions than the others. For H5N1, a greater number of AAS were observed in NA and PB2 than in the other subtypes, consistent with these segments undergoing additional selective pressure during spillover into new hosts.

## 4. Discussion

IAV-targeted probe enrichment enabled detection of multiple circulating subclades of human IAV (H1N1pdm09 and H3N2) and animal-associated H5N1 in wastewater throughout the flu season. Together, sequencing information from wastewater and clinical samples provided a more comprehensive view of circulating subtypes. The detection of H5N1 is consistent with prior research indicating that H5N1 detected in wastewater may originate from multiple sources.^29^

Notably, when wastewater IAV concentration peaks included large proportions of H5N1, corresponding peaks were not observed or were less apparent in clinical surveillance. This suggests sequencing can help users of wastewater surveillance better understand if wastewater IAV concentrations are driven by expected seasonal fluctuations of human IAV or other IAV. It also allows wastewater monitoring to capture a more comprehensive picture of circulating IAV not currently well reflected in clinical surveillance, while filling gaps arising from relying solely on M-gene PCR-based assays to interpret wastewater IAV trends. Wastewater sequencing could also be an efficient approach to track the circulation of novel variants, in particular those with markers of potential increased human adaptation. However, our ability to definitively distinguish the original host source of H5N1 in wastewater was limited for several reasons. These include the paucity of human H5N1 clinical samples available for comparison, especially of human cases residing within the same sewersheds from which wastewater samples were collected. Also, existing sequences of H5N1 circulating in cattle, avian, or human hosts can largely be indistinguishable. It is possible that, as H5N1 evolves and subtypes better suited to different hosts diverge, the ability to clearly distinguish hosts will improve. Despite limitations, our study demonstrated that probe-enriched wastewater sequencing can not only strengthen routine seasonal human influenza monitoring but also capture novel and animal-associated IAV to enhance pandemic preparedness.

This study demonstrated that it is feasible to recover multiple subtypes of IAV from wastewater with enough coverage depth to resolve strains and variants. The averaged sequencing mean coverage depth ranged from 4.5× (H5N1 PA) to 17.8× (H3N2 MP) across segments, and the mean coverage breadth ranged from 5% (H5N1 PB2) to 20.6% (H1N1 MP). A Texas surveillance study using an H5N1-focused probe panel also recovered H5N1, but the genome coverage per sample was lower.^17^ The high MP coverage aligned with previous findings, which observed a coverage bias toward smaller segments, varying considerably from longer segments.^11^ Similar to a previous H5N1 study conducted in spring 2024, we also did not find the PB2 E627K substitution for mammalian adaptation.^17^

Accurate clade deconvolution requires the capture of key mutations within antigenically important regions; however, these critical regions are frequently underrepresented in wastewater datasets. In addition, IAV sequences in the database share high similarity. Therefore, downstream bioinformatic tools might have redundant hits or random assignments that overrepresent diversity. EsViritu aimed to address this challenge by employing a read-sharing clustering algorithm to collapse redundant references to allow for the differentiation of truly different viral sequences.^16^ However, our application of EsViritu revealed a bias toward certain subtypes, such as H5N1, when we used only the conserved M-gene segment to infer subtype abundance. This limitation likely stems from the clustering process, which can obscure fine-scale genomic variation that may exist within small or conserved regions. In contrast, our EM algorithm does not perform clustering and therefore retains subtle genomic differences distributed across the genome, leading to a more accurate abundance estimation.^28^ This is specifically important in our case as we could not use the HA reads due to laboratory contamination. Adapting the EM-based tool to accommodate incomplete genomes therefore presents an opportunity to address a major challenge of inherent sequencing biases from wastewater for segmented viruses. By allowing IAV deconvolution from whatever genomic information is available, we leveraged the challenge of missing HA as an opportunity to demonstrate that EM could still extract meaningful genomic signals to resolve clade composition even when the most informative segments were absent. More work needs to be done to evaluate how accurate the proportions estimation is in comparison with the RT-PCR-based approach. Ultimately, we have shown that integrating our custom IAV probe enrichment panel with the EM algorithm provides a powerful approach for wastewater-based surveillance of IAV.

## Data Availability

The findings of this study are based on metadata associated with 100,865 sequences for building the EM reference database and the H5N1 phylogenetic tree, comprising all sequences available on GISAID up to December 27, 2025, via gisaid.org/EPI_SET_251228xr. The data for plotting Figure S1E (LACSD M-gene and H5 data during Nov-2024 to Feb-2025) were collected as part of the WastewaterSCAN / SCAN project, a partnership between Stanford University, Emory University, and Verily funded philanthropically through a gift to Stanford University. All raw sequencing data were deposited on NCBI SRA and will be publicly available upon publication.

## 5. Data sharing and code availability

The findings of this study are based on metadata associated with 100,865 sequences for building the EM reference database and the H5N1 phylogenetic tree, comprising all sequences available on GISAID up to December 27, 2025, via gisaid.org/EPI_SET_251228xr. The data for plotting **Figure S1E** (LACSD M-gene and H5 data during Nov-2024 to Feb-2025) were collected as part of the WastewaterSCAN / SCAN project, a partnership between Stanford University, Emory University, and Verily funded philanthropically through a gift to Stanford University. All raw sequencing data were deposited on NCBI SRA and will be publicly available upon publication.

## 6. Acknowledgements

We gratefully acknowledge all data contributors, i.e., the Authors and their Originating laboratories responsible for obtaining the specimens, and their Submitting laboratories for generating the genetic sequence and metadata and sharing via the GISAID Initiative, on which this research is based. We are sincerely grateful to the staff from the Los Angeles County Sanitation Districts (LACSD), Bakersfield (BKERSFLD), and Central Contra Costa Sanitary District (CCCSD) for their generous assistance with wastewater sample collection and coordination. In particular, we thank Naoko Munakata (LACSD); Salvador Lira, Hargel Pacheco, Joshua Garnica, Evette Roldan, and Kern County Department of Public Health (BKERSFLD); Blake Brown, Dan Frost, Jesse McDermott, Jason Sweet, and Lori Schectel (CCCSD), whose support made this project possible. We also thank them for permitting us to use the GIS map of their sewershed service area. We also thank California Department of Public Health’s Drinking Water and Radiation Laboratory (DWRL) staff Alexia Lopez, Bridget Hulsebosch, and Emmet Cuyler for their help with sample receiving and transfer. We prepared sequencing libraries with guidance from Casey Riegler (Twist Bioscience) and Bryan Bach (Functional Genomics Laboratory). Flow-cell loading and sequencing were performed at the Vincent J. Coates Sequencing Laboratory (QB3, UC Berkeley; RRID: SCR_022170) with advice from Shana L. McDevitt and Carrie Rose. This work was performed in part under the auspices of the U.S. Department of Energy by Lawrence Livermore National Laboratory under Contract DE-AC52-07NA27344, Release Number LLNL-JRNL-2014491. LLNL Disclaimer: This document was prepared as an account of work sponsored by an agency of the United States government. Neither the United States government nor Lawrence Livermore National Security, LLC, nor any of their employees make any warranty, expressed or implied, or assume any legal liability or responsibility for the accuracy, completeness, or usefulness of any information, apparatus, product, or process disclosed, or represent that its use would not infringe privately owned rights. Reference herein to any specific commercial product, process, or service by trade name, trademark, manufacturer, or otherwise does not necessarily constitute or imply its endorsement, recommendation, or favoring by the United States government or Lawrence Livermore National Security, LLC. The views and opinions of authors expressed herein do not necessarily state or reflect those of the United States government or Lawrence Livermore National Security, LLC, and shall not be used for advertising or product endorsement purposes.

