## Supplementary Information for "Genomic wastewater surveillance of human and animal influenza A viruses in California during the 2024-2025 flu season"

Number of Pages: 13
Number of Figures: 7
Number of Methods: 6

### Table of Contents

#### Supplementary FIgures

**Figure S1.** PMMoV-normalized M-gene and H5 concentrations across three sites
**Figure S2.** Comparison of M-gene concentrations measured by UCB dPCR and CDPH ddPCR
**Figure S3.** Fragment analysis results for (A) the pre-enriched library and (B) the enriched library
**Figure S4.** Assessment of sample contamination by tiled HA
**Figure S5.** EsViritu-estimated relative abundance of IAV clades in wastewater across three sites
**Figure S6.** Box plots of segment coverage breadth for each subtype across samples
**Figure S7.** Read coverage across IAV genome segments for a single sample (BKERSFLD, 12/12/2024)

#### Supplementary Methods

**Method A.** UCB dPCR M-gene quantification
**Method B.** CDPH DWRL methods
**Method C.** Library preparation
**Method D.** EM algorithm
**Method E.** Variant calling
**Method F.** H5N1 Phylogenetic tree
**Method G.** Equations

#### Supplementary Figures

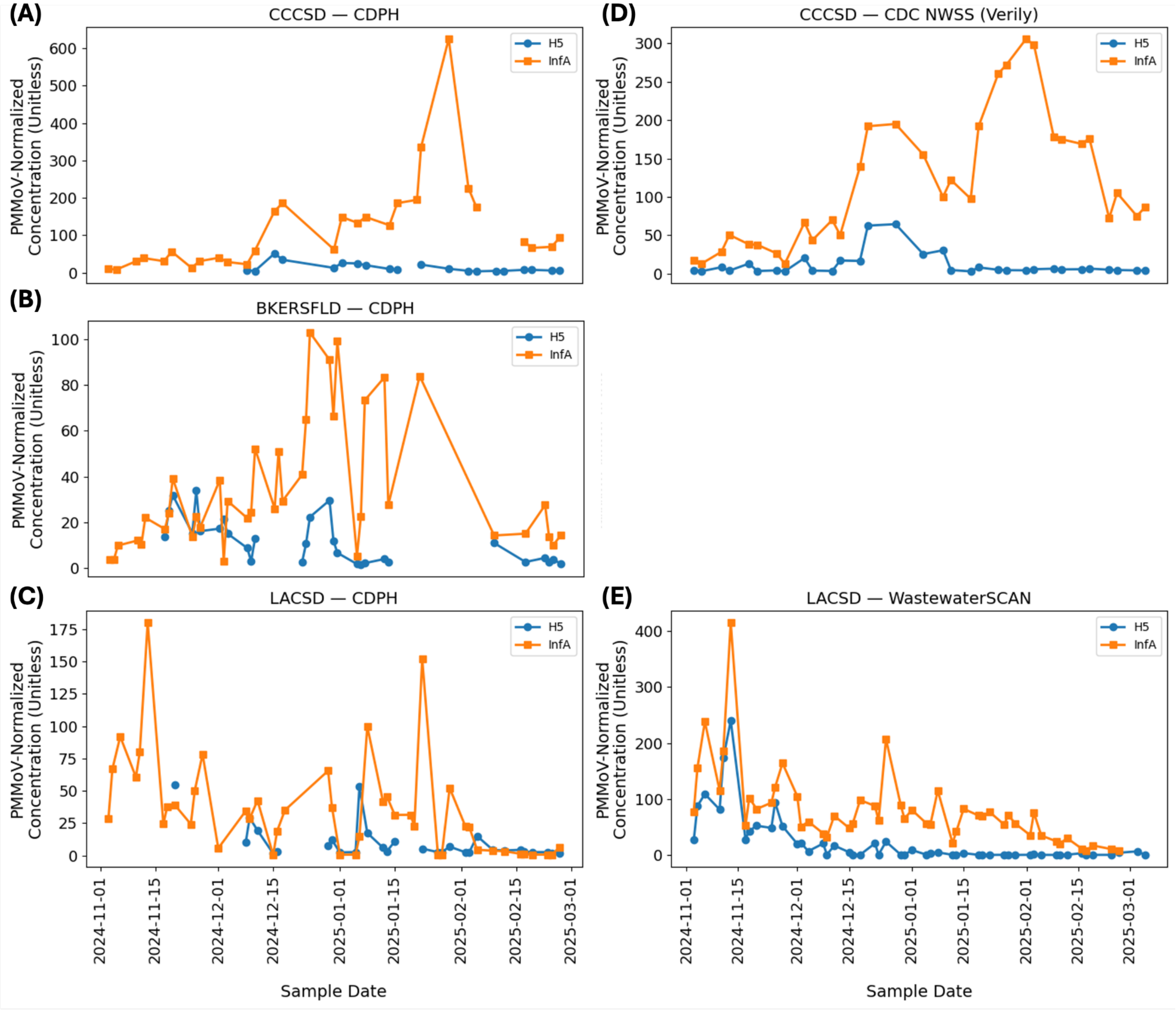


**Figure S1.** PMMoV-normalized RT-PCR M-gene and H5 concentrations (unitless) retrieved from the California Wastewater Surveillance Dashboard[1] for three sewersheds: Central Contra Costa Sanitary District (CCCSD), East Bakersfield (BKERSFLD), and Los Angeles County Sanitation Districts (LACSD), shown from top to bottom. At some sites and timepoints the IAV peaks (orange: M-gene InfA) appear to be driven by H5 (blue), while at others, total IAV is likely comprised of seasonal subtypes, however trends may be laboratory-dependent. **Panels A-C** show data generated by the California Department of Public Health (CDPH). Because H5 data from CDPH were sparse over the wastewater sampling period, data from other laboratories were included where available: CDC NWSS (Verily) for CCCSD (**Panel D**) and WastewaterSCAN for LACSD[2] (**Panel E**). No non-CDPH IAV measurements were available for BKERSFLD; therefore, only a single panel is shown for this site. All raw data used to generate this figure are provided in **Table S6**.

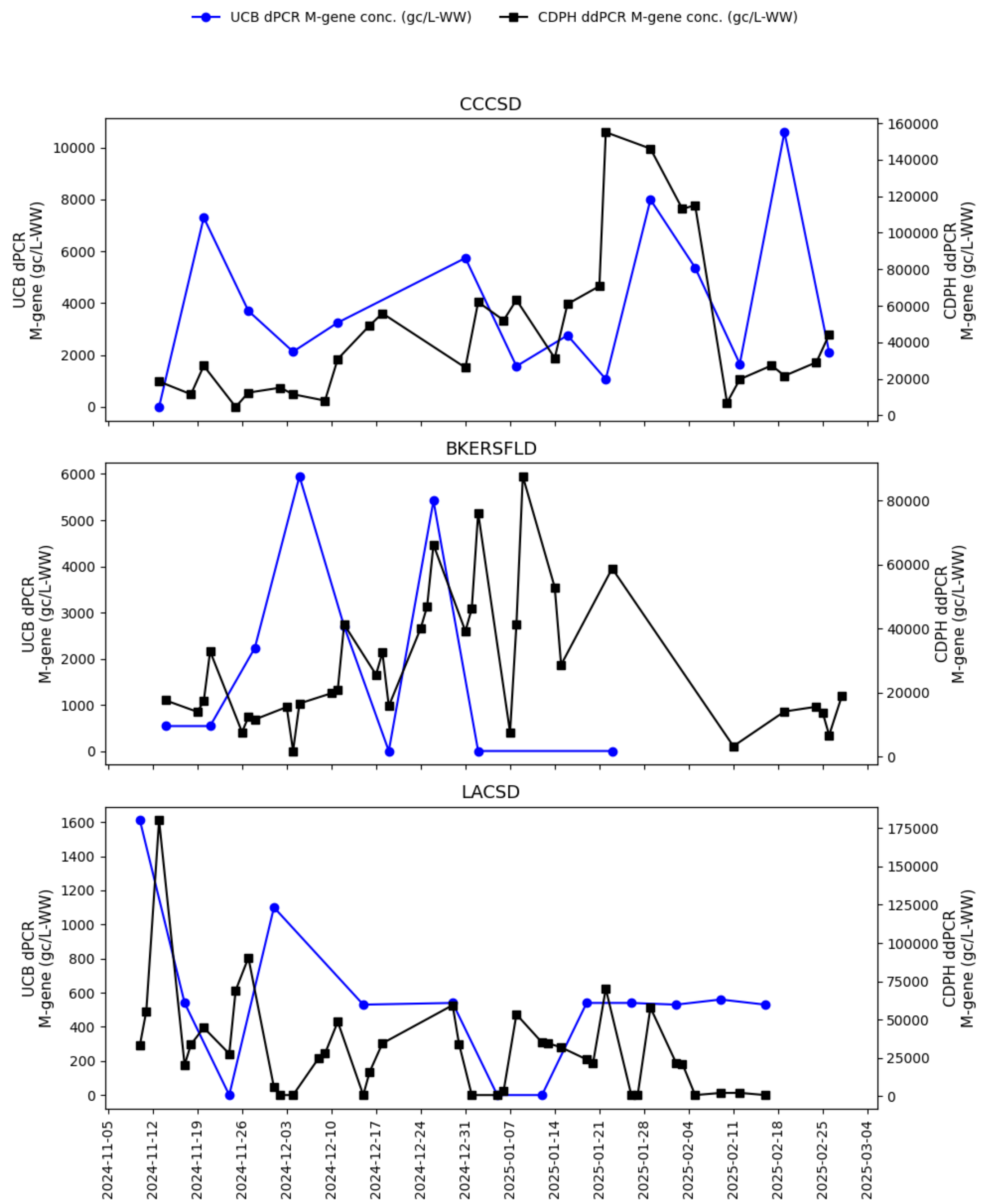


**Figure S2.** Comparison of M-gene concentrations measured by UCB (dPCR, blue line) and CDPH (ddPCR, black line). UCB samples were processed using the Promega Wizard Enviro TNA extraction protocol, and CDPH samples were processed with the Nanotrap protocol (see **Methods 2.2**). Note that some UCB measurements were having very few positive partitions, resulting in large uncertainty (wide confidence intervals).

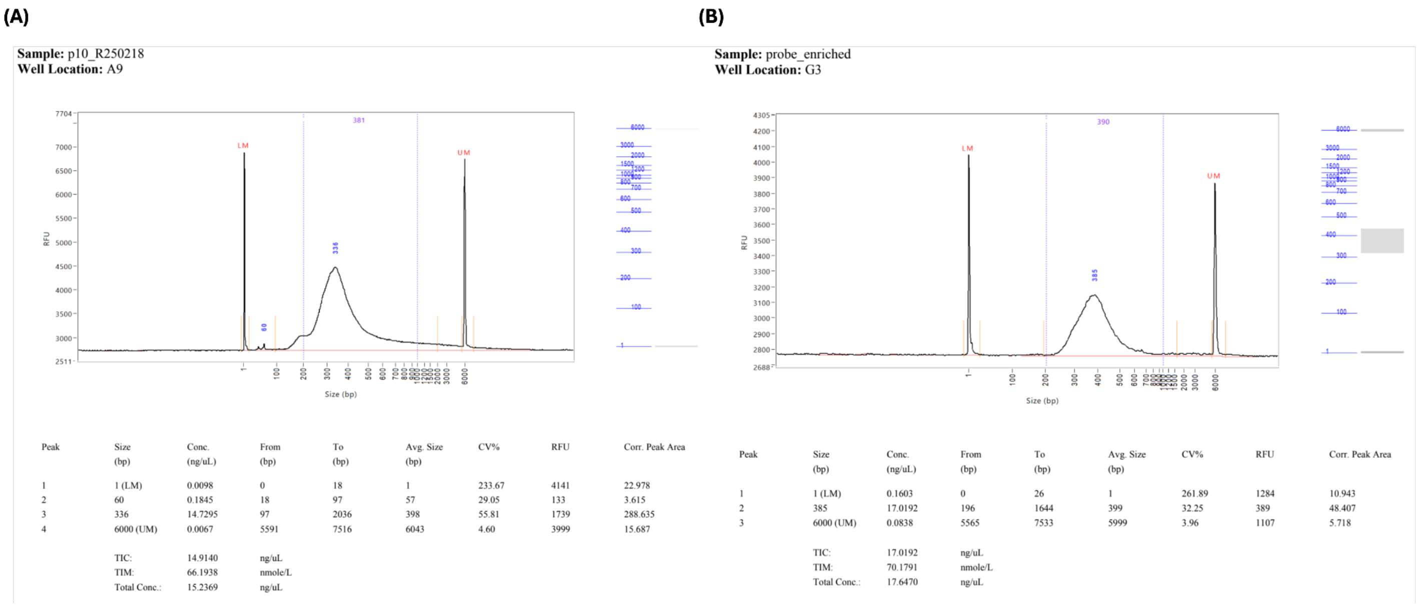


**Figure S3.** Fragment analysis results for (A) the pre-enriched library and (B) the enriched library. The pre-enriched sample shows an average fragment size of 336 bp, while the probe-enriched sample shows an average fragment size of 385 bp.

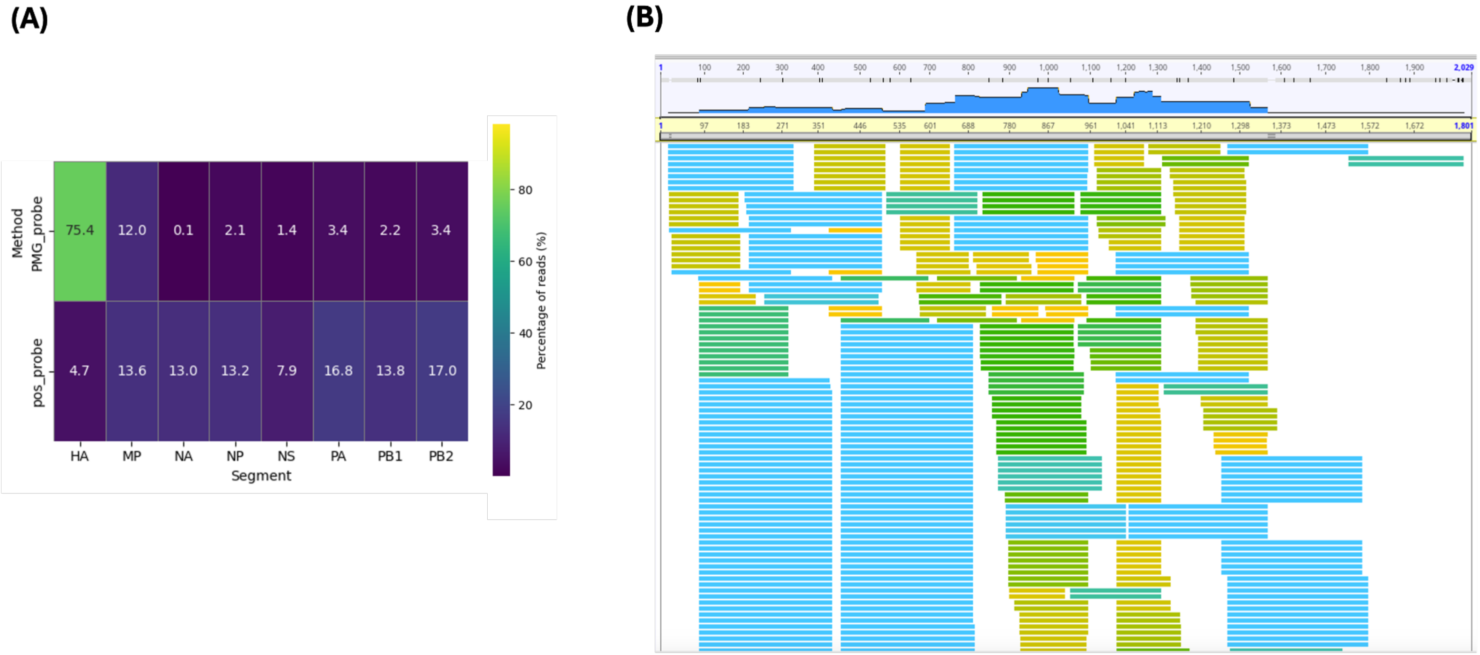


**Figure S4.** Assessment of sample contamination by tiled HA. (A) Heatmap displaying the proportion of reads aligning to each IAV segment for the Promega-extracted probe-enriched sample (PMG_probe) and for the positive control processed with IAV probe capture. (B) Coverage profile across the HA segment for the PMG_probe sample. Reads were colored by length (bp).

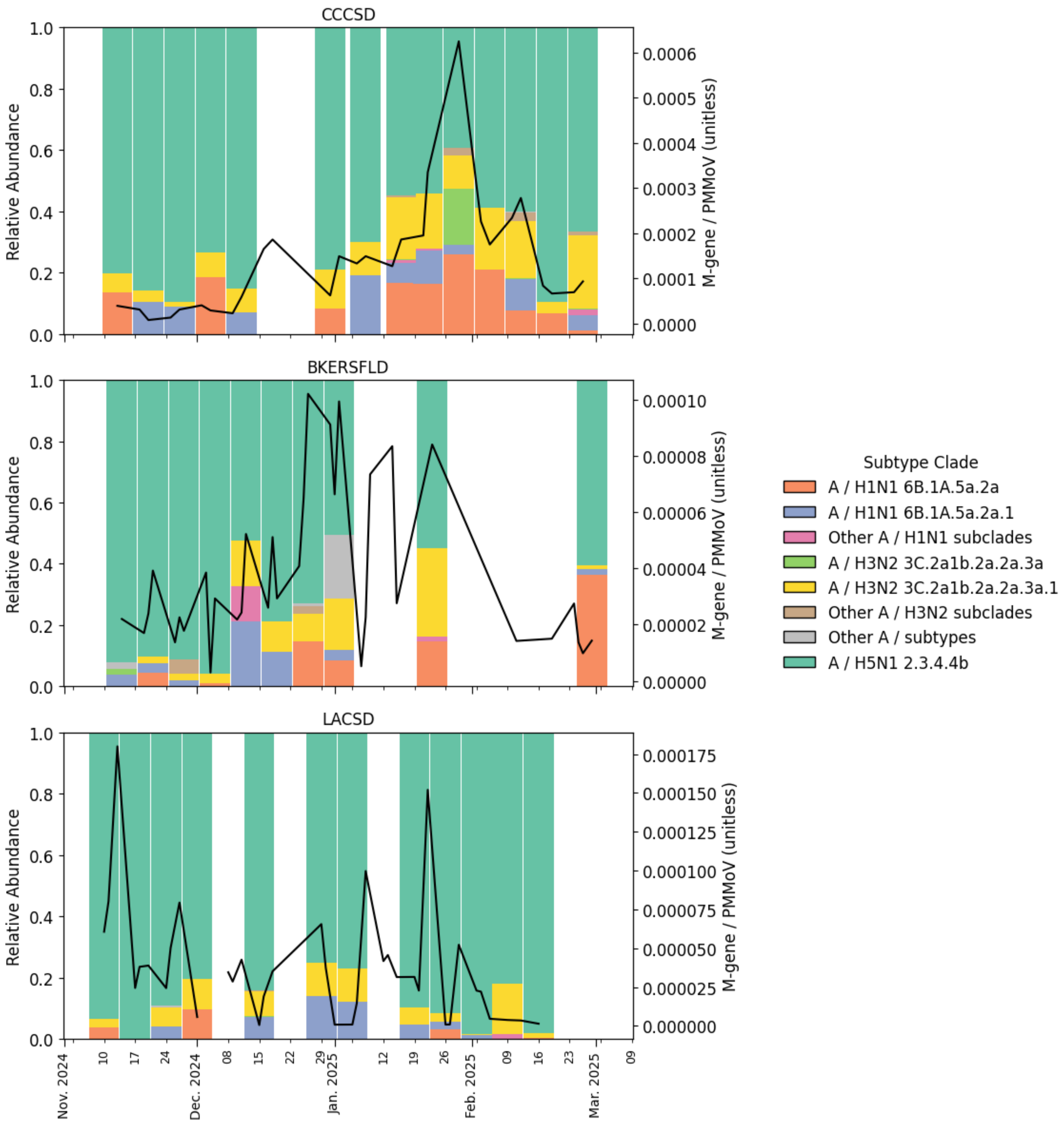


**Figure S5.** EsViritu-estimated relative abundance of IAV clades across three California wastewater treatment plants from top to bottom: Central Contra Costa Sanitary District (CCCSD, n=14), Bakersfield (BKERSFLD, n=10), and Los Angeles County Sanitation Districts (LACSD, n=12). Each stacked bar represents a weekly sequenced wastewater sample processed by IAV probe capture enrichment panel, with colors indicating different IAV clades (see legend). For each sample, clade-level relative abundance was calculated as the reads per kilobase of reference per million filtered reads (RPKMF) attributed to that clade, normalized by the total RPKMF (sum across all clades for the sample) reported in the EsViritu output. Black line indicates ddPCR PMMoV-normalized M-gene (unitless) across the sampling period.

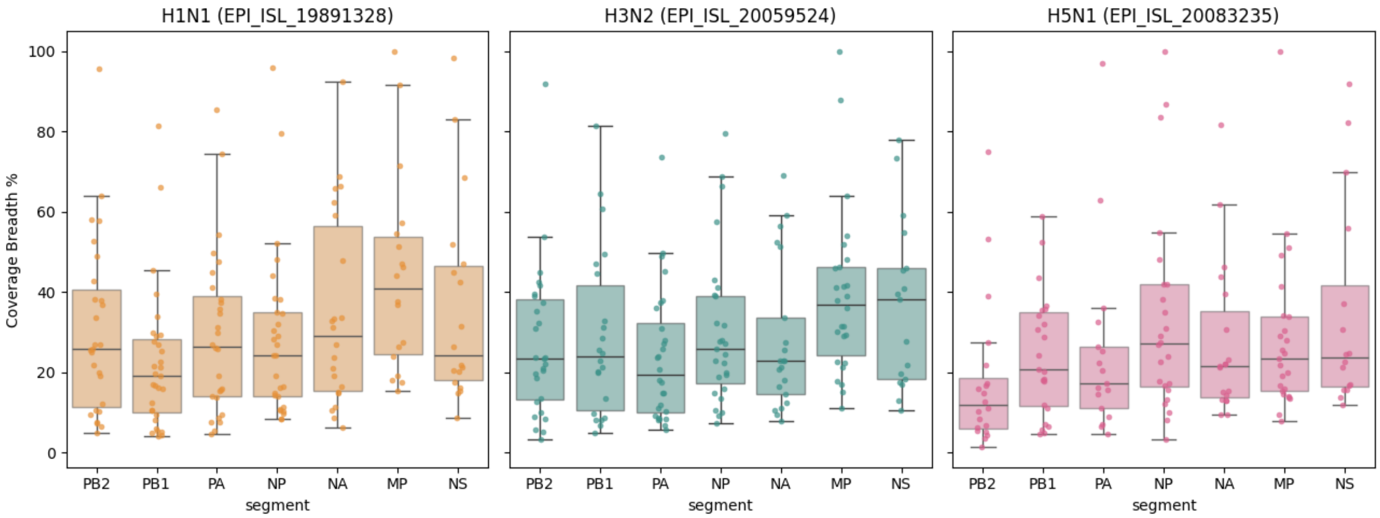


**Figure S6.** Box plots showing segment coverage breadth for each subtype across samples. Boxes and whiskers represent the interquartile range (IQR) and the minimum/maximum values within 1.5× IQR, respectively. Each dot corresponds to the segment coverage breadth (%) for a given subtype in an individual sample.

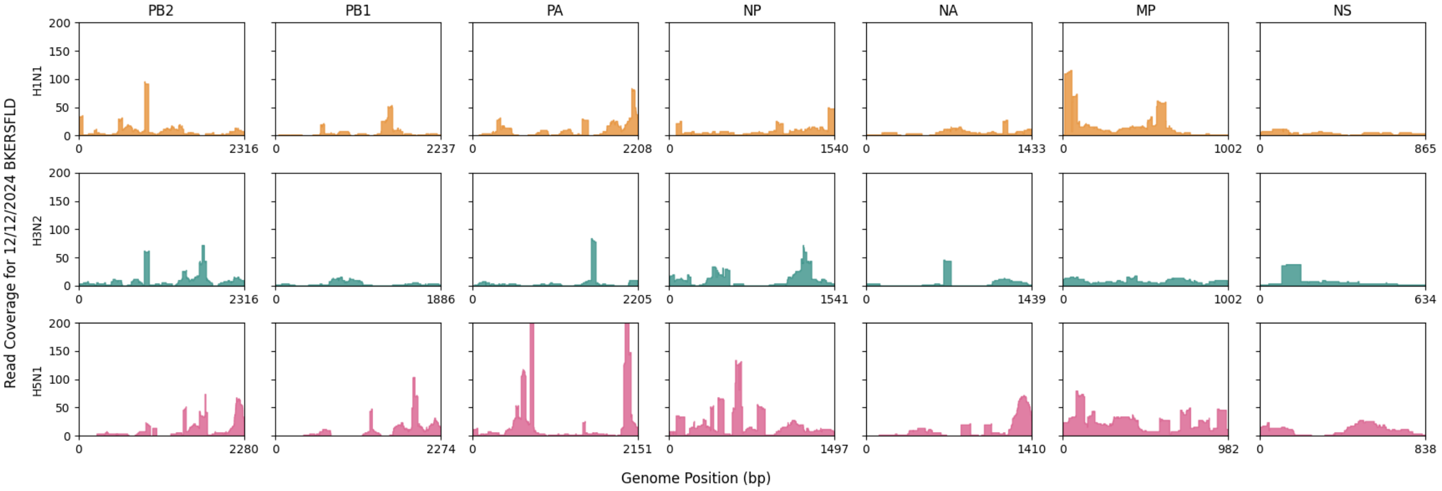


**Figure S7.** Read coverage across IAV genome segments for a single sample (BKERSFLD, 12/12/2024). Colors indicate subtypes (orange: H1N1; dark green: H3N2; dark pink: H5N1). Coverage profiles represent read counts across seven segments (excluding HA).

#### Supplementary Methods

##### A. UCB dPCR M-gene quantification

The M-gene dPCR reaction mixture (**Table S2**) was prepared using the QIAcuity OneStep Advanced Probe Kit (Qiagen). Priming and imaging were performed under default instrument settings, and thermal cycling conditions were summarized in **Table S3**. 26k nanoplates were used, with an average of 25,351 valid partitions per well and an individual partition volume of 0.2 nL. Positive controls consisted of gBlock standards (sequences shown in **Table S4**), and negative controls were nuclease-free water. All positive controls showed clear separation between positive and negative partitions, while negative controls showed zero positive partition. Below is the Environmental Microbiology Minimum Information (EMMI) Checklist[3,4] for dPCR.

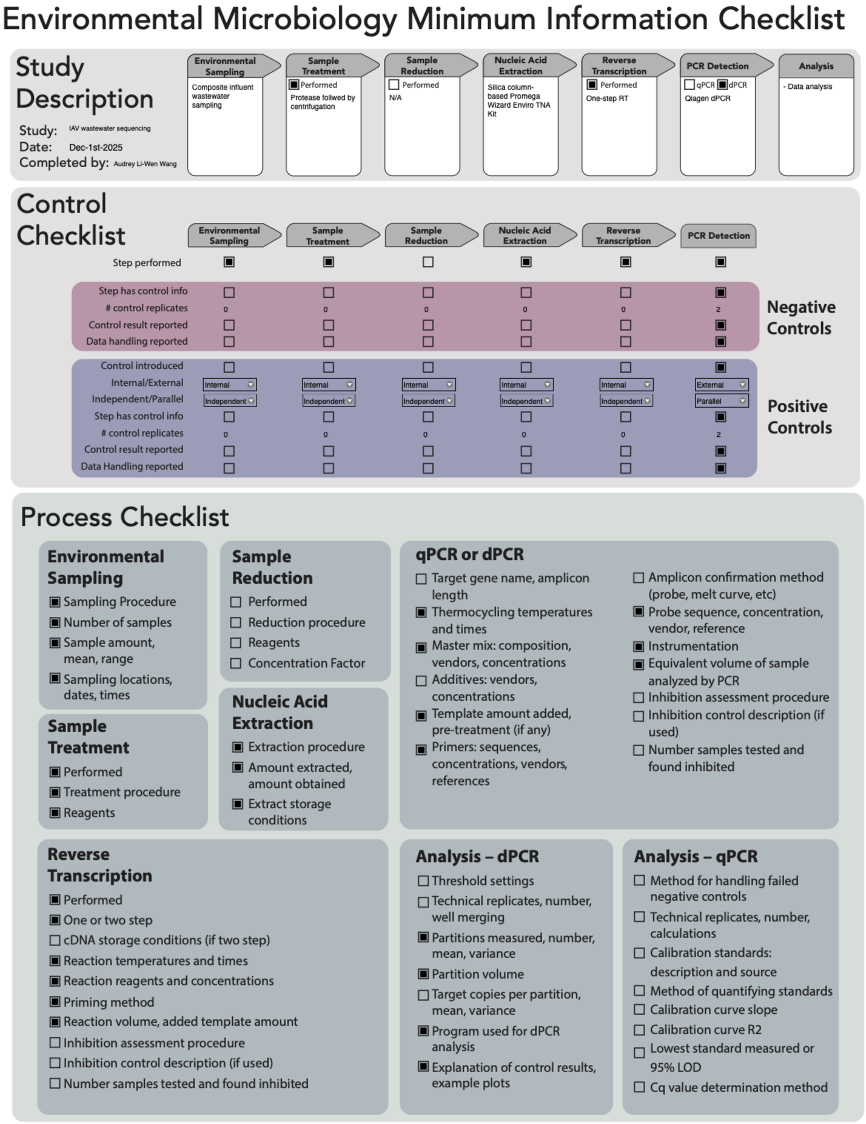

#### B. California Department of Health Drinking Water and Radiation Laboratory (DWRL) methods adapted from Karthikeyan et al. 2021[5]

##### a. Concentration and extraction methods

Influent wastewater samples were mixed by inversion followed by 10 to 15 seconds of vortexing. 5 mL aliquots of mixed wastewater influent samples were concentrated in duplicate using Ceres Nanotrap® Magnetic Virus Particles (SKU# 44202). Negative concentration/extraction controls (NEC) were prepared by conducting the concentration/extraction procedure with nuclease free water to identify cross-contamination during processing. 500 µL of the concentrated wastewater samples were eluted into MagMax Microbiome Lysis Solution (Cat# A42361) in preparation for nucleic acid extraction. Prior to extraction, lysed samples were spiked with bovine coronavirus (BCoV), as positive recovery control, to a final dilution of 1 to 1,000 (or 90 copies per microliter). The KingFisher Flex and Apex were used to extract nucleic acid (NA) from 450 µL of lysed wastewater samples using the Thermo Fisher MagMAX Viral/Pathogen Nucleic Acid Isolation Kit to a final extraction volume of 50 µL. Extracts were either analyzed immediately or stored at -80oC until analysis. Extraction duplicates were stored at -80oC for long-term storage to be used for repeating analyses in the case of quality control failures or for future analyses.

##### b. ddPCR

Nucleic acid extracts were used as a template in digital droplet RT-PCR (ddPCR) assay targeting pathogen-specific genes utilizing the primers and probes specified in **Supplementary Table S4**. Influenza M-gene was quantified as a four-plex assay together with SARS-CoV-2, respiratory syncytial virus (RSV), and mpox clade II. Influenza A H5N1 (H5 assay) was quantified as a single-plex. PMMoV was quantified as a duplex assay. The ddPCR reaction mix consisted of 5 µL of NA extract template, 5 µL of Bio-Rad One-Step RT-ddPCR Advanced Kit for Probes (CAT# 1864022), 2 µL of reverse transcriptase (CAT# 1864022), 1 µL of 300 mM dithiothreitol (DTT) (CAT# 1864022), and primer probe mixtures (PPMs) for each multiplexed pathogen target. For the multiplex assay, 1.2 µL of each of the four PPMs and 2.2 µL of nuclease-free water were added. For the H5 single plex assay, 2.4 µL of the PPM and 4.6 µL of nuclease-free water were added, and 2.4 µL of the PPMs and 2.2 µL of nuclease-free water for the duplex assay. ddPCR was performed with the following thermal cycling protocol: reverse transcription at 50°C for 60 minutes, enzyme activation at 95°C for 10 minutes, followed by 40 two-step cycles of denaturation at 94°C for 30 seconds and anneal/extension at 57°C for 1 minute. This was followed by enzyme deactivation at 98°C for 10 minutes, droplet stabilization at 4°C for 30 minutes, and hold at 6°C. Droplets were analyzed using the QX600 droplet reader (Bio-Rad) within 48 hours after PCR, and each wastewater sample was analyzed in technical triplicate utilizing the QX Manager Software 2.1 Standard Edition (Bio-Rad). Positive droplets were established utilizing the PTC and negative control wells for each individual target by setting a threshold according to the fluorescent amplitude. The thresholds were set where there was clear separation between the negative droplet baseline and the positive droplet population of the PTC wells. This threshold was then applied to all sample wells. Results from triplicate wells were merged by taking an arithmetic mean of the three wells. For a sample to be considered positive, the detected concentration should beat or above the targets' pre-determined Limits of Detection (LoDs).

#### C. Library preparation

Briefly, all samples were diluted to 3.3 ng-RNA/µL based on Qubit High Sensitivity RNA assay for a total volume of 15 µL as input for library preparation. Then, cDNA synthesis and purification by beads cleanup was performed according to the protocol. After beads cleanup, samples were diluted to 1 ng-DNA/µL for 25 µL as input cDNA for the following enzymatic fragmentation. Fragmentation time for the thermal cycler was set at 5 min to target 330 bp insert size. Fragmentation was followed by end repair and subsequent dA-tailing to generate dA-tailed DNA fragments, and each sample was ligated with Twist Universal Adapters, and after purification, each sample was indexed using Twist's unique dual index system. For barcoding PCR, 12 amplification cycles were used. QC of both DNA Qubit and fragment analysis were performed to examine the concentration and size distribution of each sample before pooling (**Table S9**). Expected average fragment length should be 250-450 bp, and we have a fragment length of 307-342 bp. An example plot of the fragment analysis results is shown in **Figure S3A**.

Eight indexed samples (also called "libraries" in the protocol) were pooled, with 187.5 ng per sample to reach a total mass of 1500 ng per pool. The library pools were dried and then hybridized with the "customized IAV probe panel" for 16 hours. Then, hybridized targets from each pool were bound to streptavidin beads, washed, and then amplified by Amplification Primers using 17 PCR cycles based on the panel size. Finally, each enriched pool was purified and examined by Qubit and fragment analyzer. The average fragment length for each enriched pool should be 375-450 bp, and our measured average fragment length falls within the range. An example plot of fragment analysis results of an enriched probe sample is shown in **Figure S3B**.

#### D. Expectation-maximization (EM) algorithm

The EM algorithm was adapted from an existing tool originally developed for SARS-CoV-2 strain deconvolution,[6] with modifications to accommodate the segmented influenza A virus genome and diverse subtypes presented in a sample. The modified implementation is available at: https://github.com/alamda/mismatch-matrix-merge. For each of three subtype anchor genomes: H1N1 (EPI_ISL_19407925), H3N2 (EPI_ISL_19407907), and H5N1 (EPI_ISL_19088566), we constructed a seven-segment "whole-genome" reference by concatenating the non-HA segments (PB2, PB1, PA, NP, NA, MP, and NS) in a fixed order. Quality-filtered reads from each sample were then aligned independently to each of the three concatenated subtype reference genomes using Bowtie2, generating three subtype-specific alignments that define the informative sites used for downstream mismatch calculations.

A seven-segment reference database consisting of 100,003 unique IAV isolates (candidate strains) was then used to evaluate agreement between reads and candidate strains. Three reference databases were used for each subtype, each containing the same set of candidate strains but differing in the subtype-specific informative sites used for calculating mismatches. Following read alignment, observed alleles at informative positions were compared against the expected alleles of each candidate strain in the reference database, and mismatch counts were computed for every read-strain pair to generate three subtype-specific mismatch matrices. For each read, mismatch counts to candidate strains were compared across the three subtype-specific matrices, and the subtype that had the lowest mismatch counts was selected to form a single consolidated mismatch matrix.

Finally, the consolidated mismatch matrix was used as input to an EM mixture model to infer the relative abundances of candidate strains within each sample. In this model, reads were treated as arising from latent candidate strains, and the likelihood of a read given a candidate strain was modeled as a function of its mismatch count, with fewer mismatches corresponding to higher likelihood. The EM algorithm iteratively updated posterior read-to-strain assignment probabilities (E-step) and strain mixture proportions (M-step) until convergence, yielding per-sample estimates of strain proportion.

#### E. Variant calling

Quality-trimmed reads were aligned to representative reference genomes for each subtype: A/California/07/2009 (H1N1), A/Darwin/9/2021 (H3N2), and A/dairy_cow/Texas/2024 (H5N1), using Bowtie2 (v2.3.4.3). Aligned reads for each sample were then processed with bcftools (v1.19) to call single-nucleotide variants (SNVs), annotate them within CDS regions, and calculate corresponding allele frequencies. Missense variant frequencies, defined as amino acid-altering mutations, were calculated using a minimum read depth of 10 and an alternative allele frequency threshold of 0.2. Amino acid changes were derived from the called variants by translating CDS-level nucleotide variants into protein-level substitutions, enabling downstream analyses of subtype-specific mutational patterns.

Suspicious reads that were assigned to special subtypes or clades were manually inspected by performing BLAST against both the NCBI and GISAID databases. These reads were found to align equally well to multiple subtypes, showing identical sequence similarity percentages. Therefore, they were classified as ambiguous and excluded from downstream analyses.

#### F. H5N1 Phylogenetic tree

All publicly available Influenza A(H5N1) sequences were downloaded from GISAID with collection dates between November 1, 2022 and November 30, 2025. In total, 19,644 viruses (157,294 segment sequences) were retrieved. Sequence data were downloaded as a multi-FASTA file, and associated metadata were obtained as a CSV file. Universal influenza A binding sites located in untranslated regions (UTRs) were removed using a custom trimming script. Each sequence underwent additional quality filtering to remove sequences containing >5% ambiguous or illegal characters or with lengths deviating by more than ±100 bp from the segment-specific median length. After filtering, 18,540 complete H5N1 genomes remained. For each virus with a complete 8-segment genome, the eight segments were concatenated in a fixed order to generate a single whole-genome sequence per isolate.

Hosts were collapsed into standardized categories (Avian, Dairy cow, Human, Swine, Other, Other mammals). Viruses annotated as Other or Other mammals were excluded, retaining only isolates from four hosts - Avian, Dairy cow, Human, and Swine hosts. After host filtering, 17,631 whole-genome sequences remained. To reduce redundancy and improve computational efficiency, whole-genome sequences were clustered using CD-HIT-EST (v4.8.1) at 99.5% nucleotide identity. We manually checked to ensure that H5n1 human clinical sequences from CDPH and our consensus wastewater H5N1 were included. The final dataset contained 2,046 whole-genome H5N1 sequences, representing avian, dairy cow, swine, human, and wastewater-associated H5N1. Multiple sequence alignment was performed using MAFFT (v7.525) with default parameters to produce an alignment of the whole-genome H5N1 sequences. A maximum likelihood phylogenetic tree was subsequently inferred using RAxML (v8.2.12) under default model settings. The resulting tree was visualized and annotated in R using the ggtree package (v4.0.1).

#### G. Equations

**IAV alignment rate (%):** The IAV alignment rate for each sample was calculated as the proportion of fastp quality-trimmed reads that aligned to the IAV reference database, where $N_{\text{IAV aligned reads}}$ is the number of reads mapped to the IAV reference database of 100,003 unique IAV isolates (excluding HA segment sequences), and $N_{\text{fastp filtered reads}}$ is the total number of reads retained after *fastp* quality trimming for that sample.

$$IAV\ alignment\ rate\ = \frac{N_{IAV\ aligned\ reads}}{N_{fastp\ filtered\ reads}} \times\ 100\% \quad \textbf{(Eq.1)}$$

**Mean depth at each genomic position (x):** For a given genomic position $i$, the mean depth was calculated as the average read depth across all samples, where $N$ is the total number of samples included in the analysis, and $d_{i,j}$ is the read depth at genomic position $i$ in sample $j$. These values were used to generate histograms of mean depth at each genomic position (**Figure 4A**).

$$d_i = \frac{1}{\mathrm{N}}\sum_{j=1}^{N} d_{i,j} \quad \textbf{(Eq.2)}$$

**Averaged segment mean depth (x):** For segment $s$ in sample $j$, the segment mean depth is shown in Eq.3, where $L_s$ is the length (bp) of segment $s$, and $d_{s,i,j}$ is the read depth at position $i$ of segment $s$ in sample $j$. The averaged segment mean depth (Eq.4) for segment $s$, averaged across $N$ number of samples is Eq.4. The values of averaged segment mean depth were used to generate the dashed line in **Figure 4A**.

$$D_{s,j} = \frac{1}{L_s}\sum_{i=1}^{L_s} d_{s,i,j} \quad \textbf{(Eq.3)}$$

$$D_s = \frac{1}{N}\sum_{j=1}^{N} D_{s,j} \quad \textbf{(Eq.4)}$$

**Coverage breadth (%):** Coverage breadth was defined as the proportion of nucleotide positions within a segment that were covered by at least one read, where $N_{\text{covered\_positions}}$ is the number of positions with sequencing depth ≥1, and L is the total length of the reference segment.

$$Coverage\ breadth\ = \frac{N_{covered\ positions}}{\mathrm{L}} \times\ 100\% \qquad \textbf{(Eq.5)}$$

**RPKMF (reads per kilobase per million filtered reads)** was calculated by normalizing the number of influenza A virus-aligned reads to the reference segment length and the total number of reads retained after *fastp* quality filtering, where $N_{\text{IAV aligned reads}}$ is the number of reads aligned to the IAV reference segment, $L_{\text{segment}}$ is the length (kp) of the reference segment, and $N_{\text{fastp filtered reads}}$ is the total number of quality-filtered reads in the sample.

$$RPKMF\ = \frac{\frac{N_{IAV\ aligned\ reads}}{L_{segment\ (kp)}}}{N_{fastp\ filtered\ reads\ (M)}} \qquad \textbf{(Eq.6)}$$

**IAV M-gene dPCR concentration (gc/L-WW):** The influenza A virus (IAV) M-gene concentration in wastewater was back-calculated based on the raw concentration measured by dPCR. $C_{\text{dPCR}}$ is the raw concentration (gc/µL) reported by the dPCR machine, $V_{\text{dPCR reaction}}$ is the final dPCR reaction volume (40 µL for a 2.6k plate), $V_{\text{dPCR template}}$ is the volume of sample template added to the dPCR reaction (10 µL for a 2.6k plate), $V_{\text{elution}}$ is the final nucleic acid elution volume (100 µL), $V_{\text{wastewater}}$ is the volume of raw wastewater processed (40-50 mL), and the factor of 1000 converts units from gc/mL-WW to gc/L-WW.

$$C_{IAV,WW}\ (gc/L) = C_{dPCR} \times \frac{V_{dPCR\ reaction}}{V_{dPCR\ template}} \times \frac{V_{elution}}{V_{wastewater}} \times 1000 \qquad \textbf{(Eq.7)}$$
